# Stratification of nuclear homogeneous patterns on HEp-2 cells based on neutrophil nuclear staining

**DOI:** 10.1101/2020.06.13.20130039

**Authors:** Dong Il Won

## Abstract

**Background:** Antinuclear antibody (ANA) testing is used to diagnose systemic autoimmune rheumatic disease (SARD). Nuclear homogeneous patterns on ANA-HEp-2 cells can result from anti-double-stranded DNA (dsDNA), anti-nucleosome, anti-histone, anti-Scl-70, or anti-dense fine speckles 70 (DFS70) antibodies (Abs). This study aimed to find a way to discriminate DFS70 Abs from others by way of assessing neutrophil nuclear staining on anti-neutrophil cytoplasmic antibody (ANCA) testing.

**Methods:** Nuclear staining on ANCA-neutrophils was assessed to stratify nuclear homogeneous patterns on ANA-HEp-2 cells. Enrolled subjects included (1) young individuals with a dense fine speckled pattern on ANA testing (young non-SARD group, n = 62) and patients with (2) systemic lupus erythematosus (SLE group, n = 33); (3) rheumatoid arthritis possibly with histone, nucleosome Abs, and others (RA group, n = 45); and (4) diffuse systemic sclerosis with Scl-70 Abs (diffuse SSc group, n = 1.4

**Results:** Negative rates (95% confidence interval) of neutrophil nuclear staining were 96.8% (88.8%– 99.6%) in the young non-SARD group, 3.0% (0.1 %–15.8%) in the SLE group, 4.4% (0.5%–15.2%) in the RA group, and 42.9% (17.7%–71.1%) in the diffuse SSc group, respectively. The negative rate of the young non-SARD group was significantly higher than those of the other groups (all *P* < 0.05).

**Conclusions:** This study suggests that the assessment of nuclear staining on ANCA-neutrophils can help to stratify nuclear homogeneous patterns on ANA-HEp-2 cells and thus to determine whether the ANA pattern is attributed to DFS70 Abs, which can be found in healthy individuals, especially in children.

## Introduction

Antinuclear antibody (ANA) testing is currently used to diagnose systemic autoimmune rheumatoid disease (SARD) [1]. This testing method is an indirect immunofluorescence (IIF) assay to assess binding of antibodies (Abs) to human HEp-2 cells that originated from human laryngeal epithelial carcinoma. The nature of the target antigen detected by SARD-associated autoantibodies differs between diseases; as such, unique reaction patterns are revealed on ANA testing. When conducting ANA testing, the fluorescence intensity (FI), reaction pattern, and Ab titer in the patient serum need to be carefully determined as the accuracy of these findings is critical to discriminate between ANA-positive healthy individuals and patients with SARD [2-4].

Meanwhile, anti-neutrophil cytoplasmic antibody (ANCA) testing is used to diagnose systemic necrotizing vasculitis or inflammatory bowel disease. The target antigens of associated Abs are constituents of the cytoplasmic granules of neutrophils, including proteinase-3 (PR3), myeloperoxidase (MPO), elastase, lactoferrin, lysozyme, and cathepsin G, among others. ANCA testing by the IIF assay employs ethanol-fixed neutrophils attached to the surface of glass slides as substrate cells [5].

Cells from the HEp-2 line are theoretically capable of infinite cell division. In contrast, circulating neutrophils, which are produced in and released from the bone marrow, do not proliferate, and typically undergo apoptosis within one day. As such, there are considerable differences observable with respect to intracellular constituents when comparing these two target cell types. As such, a comparison of the reaction patterns (ANA-HEp-2 cells and ANCA-neutrophils) may offer useful clues to help identify the target antigens of specific Abs (Fig. 1).

**Fig. 1.**
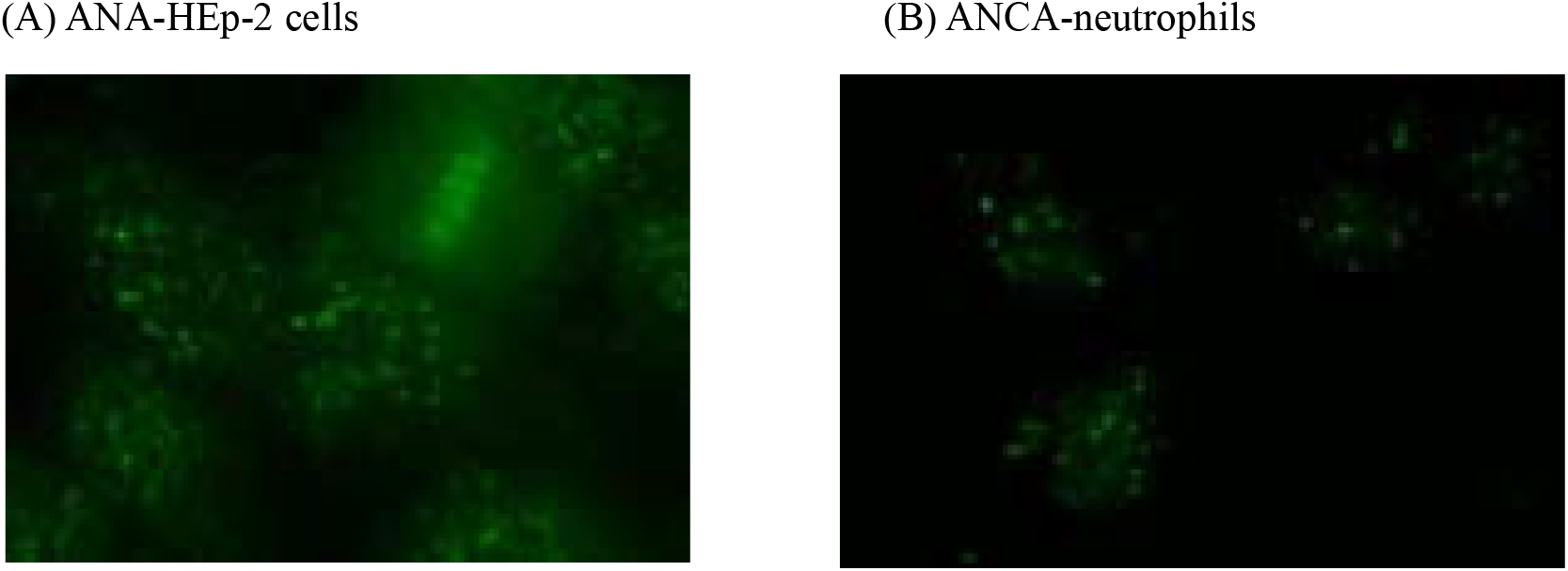
Nuclear staining patterns on (A) ANA-HEp-2 cells and (B) ANCA-neutrophils in a patient with systemic sclerosis (limited). In this case, neutrophil nuclei shared the same reaction pattern (discretely speckled) as that of HEp-2 cell nuclei. The targets of autoantibodies associated with this disease are centromeres, which are present in both substrate cells (anti-IgG-FITC staining, ×200). Abbreviations: ANA, antinuclear antibody; ANCA, anti-neutrophil cytoplasmic antibody.

Autoantibodies causing nuclear homogeneous-like patterns on ANA-HEp-2 cells include anti-double-stranded DNA (dsDNA), anti-nucleosome, anti-histone, anti-Scl-70, and anti-dense fine speckles 70 (DFS70) Abs. In details, the nuclear homogeneous pattern is characterized by smooth texture staining of the whole interphase nucleus and bright hyaline staining of the mitotic chromosomes, and the nuclear quasihomogeneous pattern is defined by an extremely fine grainy texture staining of the whole interphase nucleus with similar staining of the mitotic chromosomes. The nuclear quasihomogeneous pattern is usually associated with nucleosome Abs and/or histone Abs, whereas the nuclear homogeneous pattern is usually associated with dsDNA Abs [2]. In this report, “homogeneous-like” is used as a general term for homogeneous and quasihomogeneous.

It is well known that the DFS70 Abs can be found in healthy individuals, especially in children, whereas their clinical associations remain elusive [2,6-10]. This retrospective study aimed to discern a way by which to discriminate DFS70 Abs from other Abs when nuclear homogeneous-like patterns are observed on ANA-HEp-2 cells. For this purpose, nuclear staining on ANCA-neutrophils was paid attention to and the nuclear homogeneous-like patterns on ANA-HEp-2 cells were stratified based on this neutrophil nuclear staining in the context of ANCA testing taken together with the patient’s final diagnosis.

## Materials and Methods

ANA or ANCA testing requested by the Departments of Pediatrics or Rheumatology at Kyungpook National University Hospital (Daegu, Korea) was conducted by the Department of Laboratory Medicine. Tests were carried out by fully automated IIF assay using serum samples obtained from the patients. Nuclear staining patterns on ANCA-neutrophils were assessed when nuclear homogeneous-like patterns on ANA-HEp-2 cells were observed. The study protocol was determined to be exempt by the institutional review board of the hospital (no. KNUH 2020-01-051).

### 1. Enrolled subjects

ANA patterns were determined according to the criteria of the International Consensus on ANA Patterns (ICAP) [11]. Patients were selected by following criteria: 1) patients presented with nuclear homogeneous-like patterns on ANA-HEp-2 cells (ICAP codes AC-1, AC-2, and AC-29); and 2) patients for whom both ANA and ANCA testing were conducted simultaneously. Then, participants were classified into the following four groups according to the final diagnosis confirmed by their respective physicians: (1) individuals younger than 40 years of age not diagnosed with SARD (young non-SARD group), (2) patients diagnosed with systemic lupus erythematosus (SLE) (SLE group), (3) patients with rheumatoid arthritis (RA) (RA group), and (4) patients with diffuse systemic sclerosis (SSc) who were positive for Scl-70 Abs (diffuse SSc group). The selection criteria for each group are summarized in Table 1. In particular, the RA group included patients with Abs that targeted histones, nucleosomes, or single-stranded DNA (ssDNA) as opposed to patients with dsDNA Abs in the SLE group.

**Table 1.** Enrollment criteria and participant assignment to individual disease groups

| Disease group | Age, years | ANA testing |  |  |  | Intended target of autoantibody |
| --- | --- | --- | --- | --- | --- | --- |
|  |  | Nuclear pattern on HEp-2 cells |  | FI value, LIU | ICAP code |  |
|  |  | Interphase nucleus | Mitotic chromosome |  |  |  |
| Young non-SARD | Less than 40 | Dense fine speckled (+ few dots) | Dense fine speckled | No limit | AC-2 (+ AC-7) | DFS70 (+ p80-coilin) |
| SLE | No limit | Homogeneous | Homogeneous | Less than 1,500 | AC-1 | dsDNA |
| RA | No limit | Homogeneous | Homogeneous | Less than 1,500 | AC-1 | Histone, Nucleosome, ssDNA, etc. |
| Diffuse SSc | No limit | Homogeneous (+ speckled Nucleolus) | Homogeneous & several dots | No limit | AC-29 & AC-10 | Scl-70 (+ RNA polymerase I) |

Abbreviations: SARD, systemic autoimmune rheumatic disease; SLE, systemic lupus erythematosus; RA, rheumatoid arthritis; SSc, systemic sclerosis; ANA, antinuclear antibody; FI, fluorescence intensity; LIU, light-intensity unit; ICAP, International Consensus on ANA Patterns.

The young non-SARD group included two subgroups of patients showing (1) a dense fine speckled (DFS) pattern (ICAP code AC-2) alone or (2) a DFS pattern associated with few nuclear dots (AC-7) on ANA testing. Among them, fluorescence intensity (FI) values as determined by automated IIF were usually measured at levels less than 1,500 light intensity units (LIU). As such, an upper limit (1,500 LIU) was applied to the FI value when selecting among the patients with SLE or RA so that the FI values of these groups would not differ significantly from those of the young non-SARD group. When selecting cases from among those with diffuse SSc, patients were included only if speckled nucleoli and several dots on mitotic chromosomes (AC-10) were detected together with the nuclear homogeneous patterns on ANA-HEp-2 cells (AC-29).

The characteristics of the participants selected and assigned to each category are presented in Table 2. As intended, FI values on ANA testing of the SLE or RA groups showed no significant difference relative to those of the young non-SARD group (unpaired *t*-test).

**Table 2.** Demographics of enrolled subjects according to each disease group

| Disease group | n | Sex (M:F) | Age, years (range) | FI on ANA testing |  | Positive rate of disease-marker antibody |
| --- | --- | --- | --- | --- | --- | --- |
|  |  |  |  | FI value | P-value* |  |
| Young non-SARD | 62 | 22:40 | 18.0 (2–38) | 503.5 ± 476.3 |  |  |
| SLE | 33 | 1:32 | 37.3 (13–72) | 590.1 ± 468.8 | 0.398 | dsDNA Ab: 93.9% (31/33) |
| RA | 45 | 10:35 | 59.5 (24–83) | 460.0 ± 500.4 | 0.649 |  |
| Diffuse SSc | 14 | 2:12 | 53.9 (34–69) | 1,568.1 ± 652.7 | < 0.05 | Scl-70 Ab: 100% (14/14) |

\*FI values of the non-SARD group were compared with those of the other disease groups (unpaired *t*-test).
Abbreviations: SARD, systemic autoimmune rheumatic disease; SLE, systemic lupus erythematosus; RA, rheumatoid arthritis; SSc, systemic sclerosis; ANA, antinuclear antibody; FI, fluorescence intensity; Ab, antibody.

### 2. Automated IIF testing

ANA or ANCA slide processing, from specimen dilution to the final wash steps, was conducted using a QUANTA-Lyser instrument. Digital images of stained HEp-2 cells or neutrophils were acquired using the NOVA View instrument with accompanying version 2.0 software. Two independent experts reviewed these images on liquid crystal display monitors using the QUANTA Link (all from Werfen, Barcelona, Spain) [12].

NOVA Lite HEp-2 IgG ANA with DAPI kits were used for ANA screening with sera diluted at 1:80, while NOVA Lite ANCA (ethanol-fixed) kits were used for ANCA screening with sera diluted at 1:20 (both from Werfen). The cutoff values used to discriminate between positive and negative FI values were 48 LIU for ANA testing and 20 LIU for ANCA testing, respectively.

Nuclear staining on ANCA-neutrophils was assessed to stratify nuclear homogeneous-like patterns on ANA-HEp-2 cells. For this purpose, solid nuclear (e.g., Fig. 2B, showing an SLE case), or intranuclear patterns (e.g., Fig. 2D, showing an SSc case) on ANCA-neutrophils were assigned as positive neutrophil nuclear staining. Besides, very perinuclear (atypical P-ANCA) patterns accompanied by these intranuclear patterns (e.g., Fig. 2C, showing an RA case) were also assigned as indicating positive neutrophil nuclear staining. Like these, the interpretation criteria of neutrophil nuclear staining in the current study were different from those of conventional ANCA testing to diagnose systemic necrotizing vasculitis or inflammatory bowel disease.

**Fig. 2.**
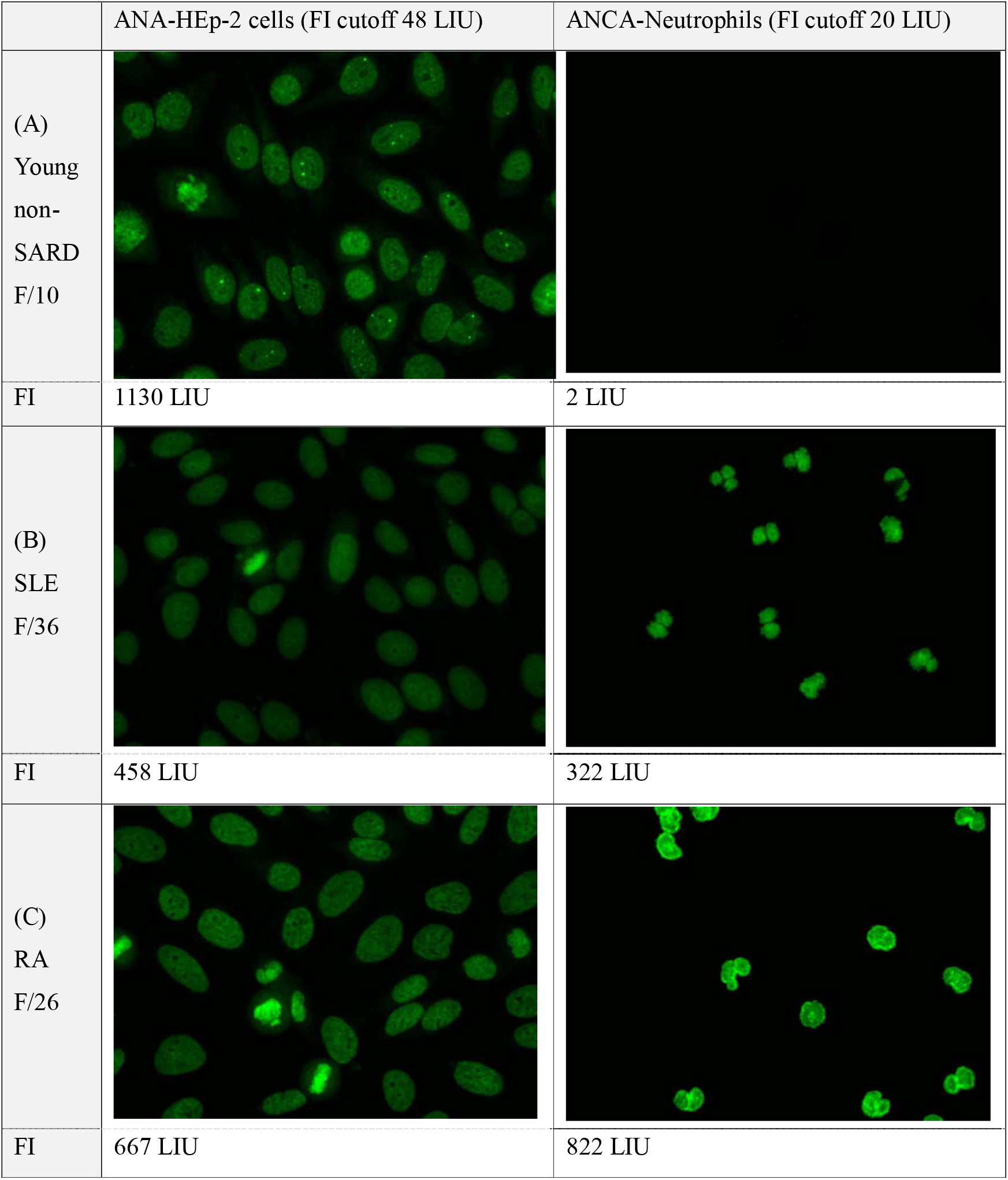
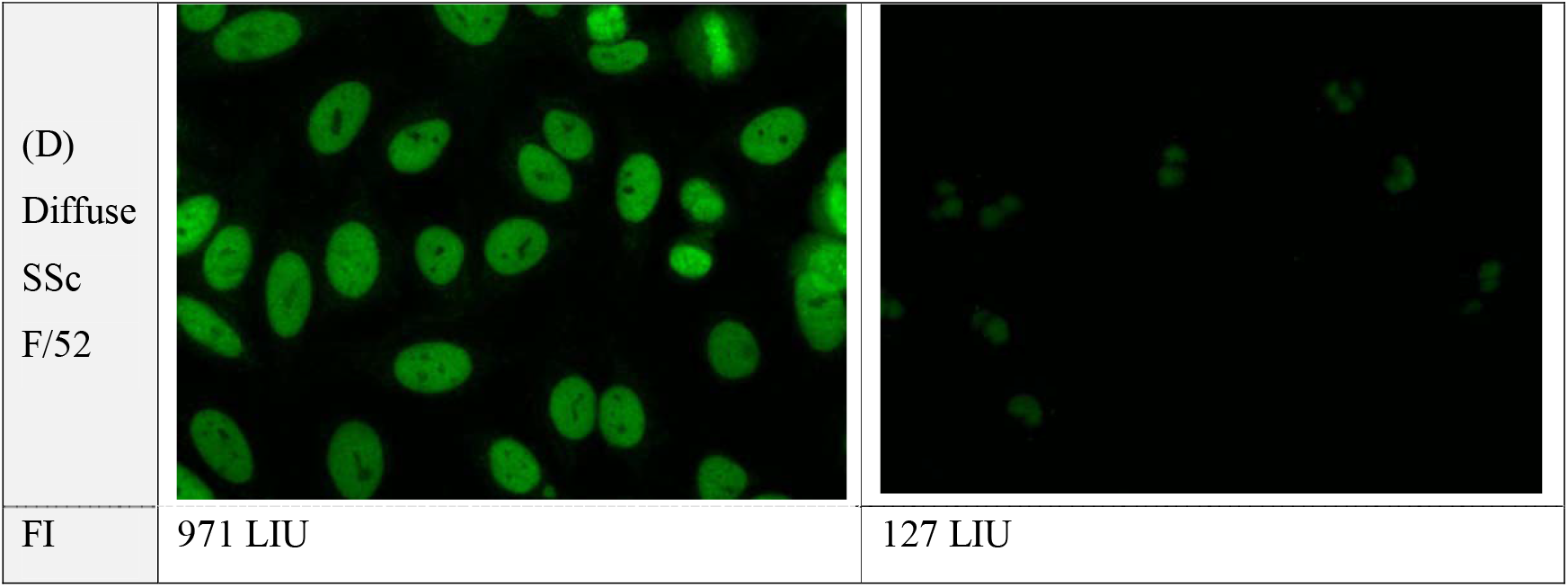
Nuclear staining patterns of HEp-2 cells and neutrophils in patients with nuclear homogeneous-like patterns on ANA-HEp-2 cells. The staining of the two cell types was performed in the context of ANA testing and ANCA testing, respectively. A representative case for each disease group is presented. In HEp-2 cell nuclei, the case of a young individual (A) who was not diagnosed with SARD (non-SARD) shows a dense fine speckled pattern (AC-2) as well as a pattern of few nuclear dots (AC-7). The case of diffuse SSc (D) presents both a homogeneous pattern (AC-29) and a speckled nucleolar pattern with several dots visible on mitotic chromosomes (AC-10). With respect to nuclear staining on ANCA-neutrophils, the young non-SARD case (A) shows negative staining, whereas the other cases (B–D) show positive staining (anti-IgG-FITC staining, ×200). Abbreviations: SARD, systemic autoimmune rheumatic disease; SLE, systemic lupus erythematosus; RA, rheumatoid arthritis; SSc, systemic sclerosis; ANA, antinuclear antibody; ANCA, anti-neutrophil cytoplasmic antibody; FI, fluorescence intensity; LIU, light intensity unit.

### 3. Statistical analysis

Statistical analyses were performed using the Statistical Package for the Social Sciences version 23.0 software program (IBM Corporation, Armonk, NY, USA). An unpaired *t*-test was used to compare FI values between two groups. For negative rates of nuclear staining on ANCA-neutrophils, 95% confidence intervals were obtained and patient groups were compared with Fisher’s exact test. *P*-values of less than 0.05 were considered to be statistically significant. Data are expressed as mean ± standard deviation or range (minimum–maximum).

## Results

Nuclear homogeneous-like patterns on ANA-HEp-2 cells were stratified based on nuclear staining on ANCA-neutrophils according to the four disease groups.

### 1. Fluorescent microscopic images of ANA and ANCA testing

Representative fluorescent microscopic images of both ANA-HEp-2 cells and ANCA-neutrophils for each group with nuclear homogeneous-like patterns on ANA-HEp-2 cells are presented in Fig. 2. Regarding nuclear staining on ANCA-neutrophils in these representative images, the young non-SARD case shows negative staining, whereas the other cases show variable positive staining as follows: solid nuclear patterns (in Fig. 2B, an SLE case), intranuclear patterns (in Fig. 2D, an SSc case), and very perinuclear patterns (atypical P-ANCA) accompanied by intranuclear patterns (in Fig. 2C, an RA case).

### 2. Changes in FI (HEp-2 cells *vs*. neutrophils) in a single patient according to each group

Among patients with nuclear homogeneous-like patterns on ANA-HEp-2 cells, changes in FI were evaluated (HEp-2 cells *vs*. neutrophils, Fig. 3).

**Fig. 3.**
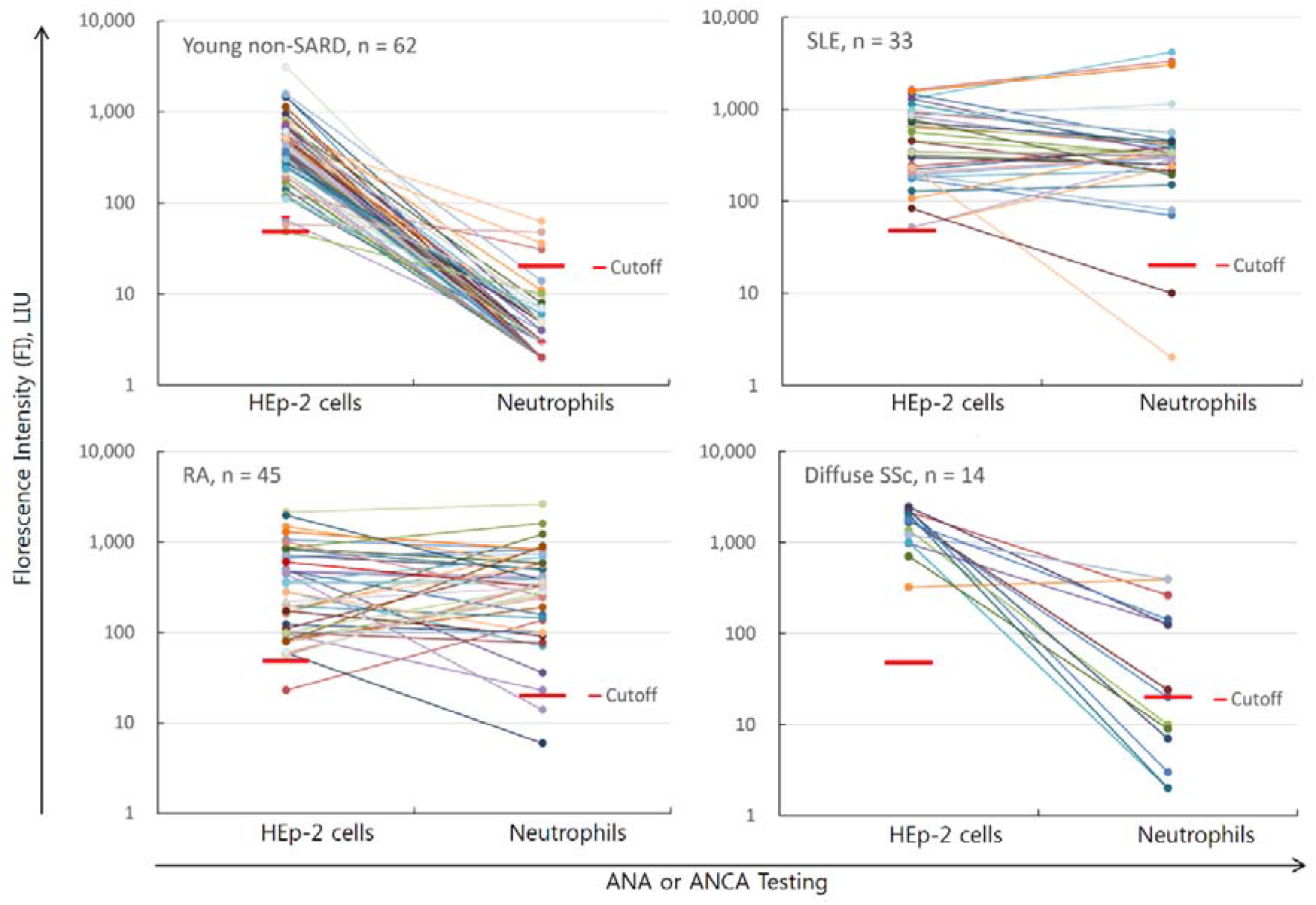
Changes in FI (HEp-2 cells *vs*. neutrophils) in a single patient with nuclear homogeneous-like patterns on ANA-HEp-2 cells. The changes are presented according to each disease group (i.e., young non-SARD, SLE, RA, and diffuse SSc). Most patients in the young non-SARD group showed an abrupt decline in FI values. The results of all three group pairings for the comparison of FI values on ANCA testing with adopting the young non-SARD group as the reference group were significant (*P* < 0.05, unpaired *t*-test). Abbreviations: SARD, systemic autoimmune rheumatic disease; SLE, systemic lupus erythematosus; RA, rheumatoid arthritis; SSc, systemic sclerosis; FI, fluorescence intensity; LIU, light intensity unit; ANA, antinuclear antibody; ANCA, anti-neutrophil cytoplasmic antibody.

### 3. Qualitative assessment of neutrophil nuclear staining or FI values on ANCA testing

Among patients with nuclear homogeneous-like patterns on ANA-HEp-2 cells, statistical analysis of the qualitative (visual determination of neutrophil nuclear staining) and quantitative results (measured FI values) on ANCA testing is presented in Table 3.

**Table 3.**
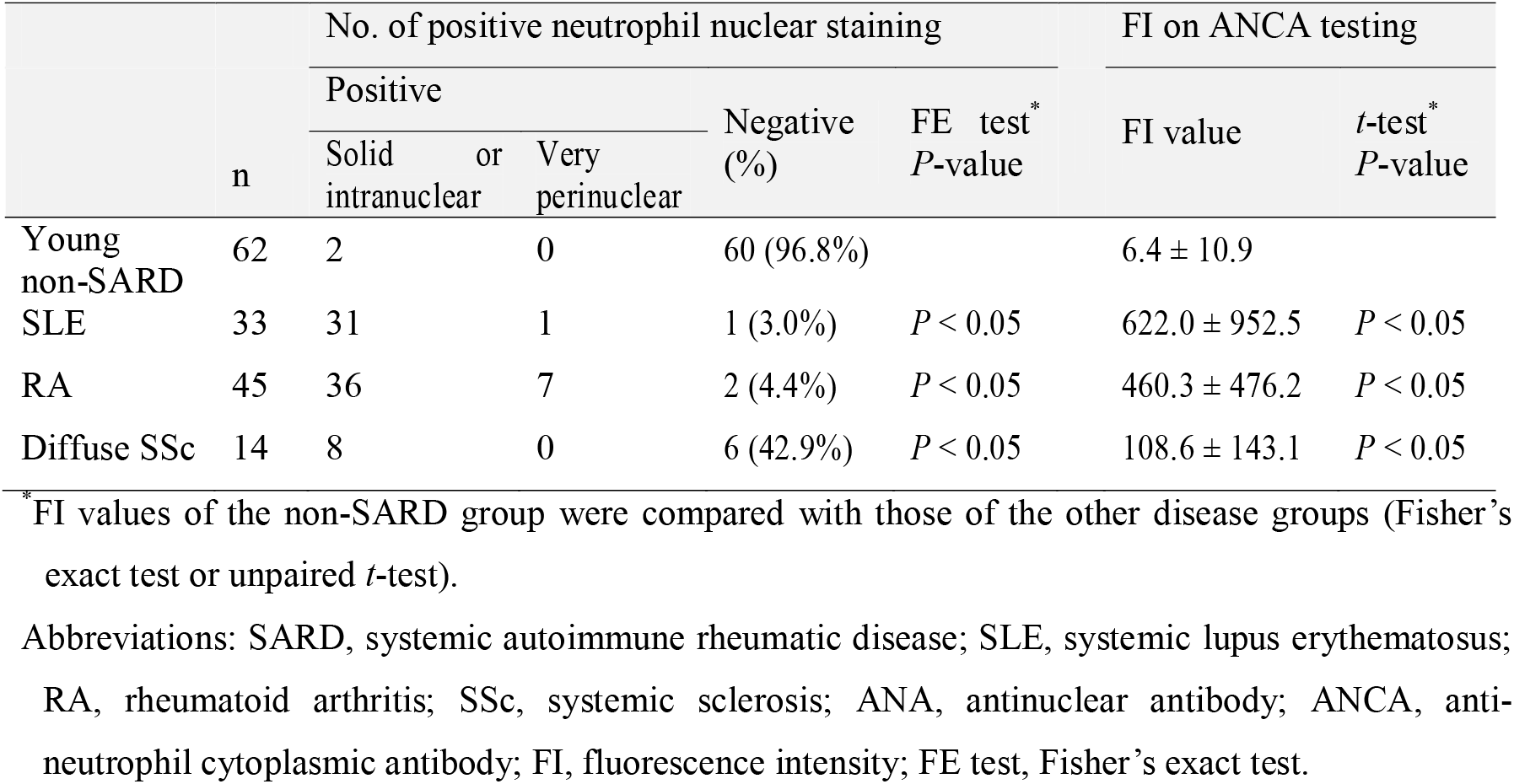
Qualitative and quantitative results on ANCA testing according to each disease group with the nuclear homogeneous-like pattern on ANA-HEp-2 cells.

The negative rates of nuclear staining on ANCA-neutrophils were 96.8% (88.8%–99.6%) in the young non-SARD group, 3.0% (0.1 %–15.8%) in the SLE group, 4.4% (0.5%–15.2%) in the RA group, and 42.9% (17.7%–71.7%) in the diffuse SSc group. Regarding positive to negative qualitative determinations, the difference between the two disease groups in all three group pairings was significant (*P* < 0.05, Fisher’s exact test) when using the young non-SARD group as the reference group. The comparison of FI values following ANCA testing of all three pairs was also significant (*P* < 0.05, unpaired *t*-test).

In this study, the eight ANCA tests that revealed very perinuclear patterns were also regarded as positive neutrophil nuclear staining given that all cases were accompanied by solid nuclear or intranuclear patterns (see Fig. 2C, showing an RA case). In two cases associated with the young non-SARD group, FI values on ANCA testing exceeded the cutoff value of 20 LIU at 31.0 and 63.0 LIU, respectively, although the nuclear staining on ANCA-neutrophils was visually negative in both cases. Both cases seemed to exhibit an increase in FI values due to nonspecific staining of intercellular background areas (i.e., false-positive FI values). Two of the cases assigned to the SLE group also had no positive neutrophil nuclear staining. In both cases, the levels of dsDNA Abs in serum were also negative (4.07 and 1.25 IU/mL, reference value < 7.0 IU/mL).

## Discussion

Pediatricians often request ANA testing to rule out SARD in an otherwise healthy child with microscopic hematuria of unknown origin or skin rash. Differentiating nuclear homogeneous-like patterns on ANA-HEp-2 cells is not straightforward based on ANA testing alone. A laboratory report that misreads a DFS pattern as a homogeneous one can trigger unnecessary and unwarranted anxiety. At this time, the physician should assess the clinical importance of this result based on secondary testing, such as ANA panel testing for dsDNA, DFS70, or extractable nuclear antigens (ENA; e.g., Sm, RNP, SSA/Ro, SSB/La, Scl-70, Jo-1, and CENP-B, among others). However, performing all these examinations is not efficient.

This study suggests that the assessment of nuclear staining on ANCA-neutrophils can help to differentiate anti-DFS70 Abs from anti-dsDNA, anti-nucleosome, or anti-histone Abs in patients who initially present with nuclear homogeneous-like patterns on ANA-HEp-2 cells: The nuclear staining on ANCA-neutrophils is routinely negative for DFS70 Abs and positive for Abs targeting dsDNA, histones, and nucleosomes.

The diseases associated with nuclear homogeneous-like patterns on ANA testing include (1) SLE positive for dsDNA Abs; (2) SLE, RA, or autoimmune hepatitis (AIH) positive for nucleosome Abs; and (3) diffuse SSc positive for Scl-70 Abs. The dsDNA Abs are unique to patients with SLE and are detected in 75% to 90% of patients with active disease. Meanwhile, nucleosome Abs are detected in 75% of all SLE patients, in 100% of those diagnosed with drug-induced lupus, and in 20% to 50% of patients with AIH type I [11].

In a previous study that explored the target specificity of autoantibodies in 59 ANA-positive RA patients, histone Abs were detected most often (14/59) followed by Abs targeting ssDNA (4/59), SS-A (4/59), and dsDNA (2/59), respectively [13]. RA patients treated with anti-TNFα (infliximab) may gradually develop dsDNA Abs and nucleosome Abs for a period of 30 weeks [14]. In RA patients, very perinuclear patterns are frequently observed on ANCA testing as the result of autoantibodies targeting lactoferrin, among others [15]. In the current study focusing on nuclear homogeneous-like patterns on ANA-HEp-2 cells, very perinuclear patterns on ANCA testing were observed in seven out of 45 RA patients and were always accompanied by these intranuclear patterns on ANCA-neutrophils (Fig. 2C. showing an RA case), i.e., positive neutrophil nuclear staining.

The Scl-70 Ab targets Scl-70 (DNA topoisomerase I), which is detected in patients with diffuse SSc and shows a nuclear homogeneous pattern on ANA-HEp-2 cells (AC-29). This homogeneous pattern per se is indistinguishable from that triggered due to the presence of dsDNA Abs [16]. The additional findings of speckled nucleoli and several dots on the positive mitotic chromosomes of ANA-HEp-2 cells (AC-10) can help to differentiate Scl-70 Abs from dsDNA Abs. This unique additional pattern is linked to Ab targeting of RNA polymerase I and/or nucleolar organizing regions (NOR)-90 (hUBF) [17]. It should be noted that the Scl-70 Abs identified in patients with diffuse SSc (AC-29) may lead to negative nuclear staining on ANCA-neutrophils. Fortunately, it is possible to differentiate Scl-70 Abs from others based on the aforementioned additional ANA findings alone. In the current study, solid or intranuclear staining on ANCA-neutrophils was found in seven of the 13 cases of the diffuse SSc group; this appears to be due to the presence of nucleosome, histone, or ssDNA Abs, among others, accompanied by Scl-70 Abs.

The DFS70 Ab shows the DFS pattern (AC-2) on both interphase nuclei and mitotic chromosomes of HEp-2 cells. The target antigen of this autoantibody is DFS70, a protein also known as the DNA binding transcription coactivator p75 or lens epithelium-derived growth factor [6,7]. In healthy children, this reaction pattern is often accompanied by one to six nuclear dots (p80-coilin Abs, AC-7) [8]. Of the 62 individuals in the young non-SARD group (younger than 40 years of age) who showed DFS patterns, 16 individuals also presented this pattern (few nuclear dots) in the current study. The finding of this nuclear DFS pattern on HEp-2 cells can problematic because it can be difficult to distinguish from nuclear homogeneous patterns due to the other autoantibodies associated with SARD.

Carbone *et al*. [18] conducted 14 tests for disease-marker autoantibodies in a total of DFS70 Ab-positive 55 adults (children were not included among the study subjects). According to this study, all men (10/55) tested negative. Among the women (45/55), about half of the women (23/45) tested positive. Anti-thyroid peroxidase Ab was the most common of the Abs detected (7/23); moreover, one subject tested positive for Ab against Mi-2, a member of the ENA family, and three subjects were also ANCA-positive, with Abs targeting MPO, PR3, or both. Concerning the clinical features of DFS70 Ab-positive subjects, the majority of subjects (37/55) were healthy, while the remaining subjects (18/55) were diagnosed with various diseases including four subjects with celiac disease. This study suggested the presence of a DFS pattern in adults should trigger a follow-up investigation for autoimmune diseases, especially among women. Nevertheless, the DFS70 Abs detected in adults may be more or an epiphenomenon associated with immune system dysregulation and this Ab per se may not play a pathogenic role.

Antigens associated with nuclear homogeneous-like patterns on ANA-HEp-2 cells such as dsDNA, ssDNA, nucleosome, histone, and DFS70 may be rich in HEp-2 cell nuclei. However, DFS70 and Scl-70 antigens appear to be absent in neutrophil nuclei. This is the basis of the proposed strategy used to stratify nuclear homogeneous-like patterns on ANA-HEp-2 cells in the current study.

In the current study, the specificities of the following two kinds of autoantibodies were not confirmed by a solid phase assay: DFS70 Abs in the young non-SARD group and histone Abs in the RA group. This limitation seems to have been overcome with close observation of the nuclear reaction patterns on ANA-HEp-2 cells. However, there are other limitations to the current study. First, patients diagnosed with AIH were not included in this study due to their limited numbers. Nuclear homogeneous patterns on ANA-HEp-2 cells can be observed in AIH patients due to nucleosome Abs [19]. Second, the ANA or ANCA slides used were from a single company (Werfen). The specific nature of the fixative solution and process used to pretreat HEp-2 cells and neutrophils may result in the denaturation or dissolution of relevant cellular antigens [20]. As such, further studies are required to determine whether the results of the current study can be reproduced using slides from other manufacturers. Finally, further studies confirming that DFS70 and Scl-70 antigens are not detectable in neutrophil nuclear extracts by immunoblotting would offer key support for the current study.

In conclusion, this study suggests the assessment of nuclear staining on ANCA-neutrophils can help to stratify nuclear homogeneous-like patterns on ANA-HEp-2 cells and thus to determine whether ANA patterns are attributed to DFS70 Abs, which can be found in healthy individuals, especially in children.

## Data Availability

.

